# Dental-Facial Midline: An Esthetic Based Classification

**DOI:** 10.1101/2021.04.24.21255904

**Authors:** Nischal Niraula, Reecha Acharya, Manoj Humagain, Zohaib Khurshid, Necdet Adanir, Dinesh Rokaya

**Affiliations:** Department of Periodontology and Implantology, Kathmandu University School of Medical Sciences, Dhulikhel, Kavre, Nepal; Department of Public Health, Torrens University Australia, Sydney, Australia; Centre for Oral Health Outcomes & Research Translation, Campbell street Liverpool NSW 2170, Australia; Ingham Institute for Applied Medical Research, Liverpool BC, NSW 1871, Australia; Western Sydney University, School of Nursing and Midwifery, Rydalmere NSW 2116, Australia; Department of Prosthodontics and Dental Implantology, School of Dentistry, King Faisal University, Al-Ahsa 31982, Saudi Arabia; Department of Restorative Dentistry, College of Dentistry, King Faisal University, Al Ahsa, Saudi Arabia; Department of Clinical Dentistry, Walailak University International College of Dentistry, Walailak University, Bangkok, Thailand

**Keywords:** Esthetics, Face, Midline, Dental, Asian, Nepalese

## Abstract

**Background:** The facial midline and dental midline play an important role in facial esthetics, cosmetic dentistry, facial plastic surgery, and anthropologic studies.

**Objectives:** This study studied the dental-facial midline in Nepalese subjects and to classify the midline.

**Methods:** The study was done in 150 Nepalese subjects, mostly consisting of University students (80 males and 70 females). After obtaining ethical approval, facial and dental midlines were analyzed using a scale.

**Results:** It showed 26 (18%) study subjects showed the coincidence of the facial midline with the maxillary and mandibular dental midlines. It showed that only 44 (30%) subjects showed the coincidence of facial midline with only maxillary dental midline, and 26 (17%) subjects showed the facial midline coincidence with only mandibular dental midline. The dental midline discrepancy was more prevalent in the maxillary arch and more prevalent on the right side. Midline discrepancy is seen more in males compared to females. The majority of the deviation showed 1 mm, followed by 2 mm, and 3 mm.

**Conclusions:** The coincidence of the facial midline with both the maxillary and mandibular dental midlines is uncommon. Midline discrepancy is seen more in males compared to females. The majority of the subjects show a mild discrepancy of 1 mm. The midline discrepancy was more seen on the right side and in the maxillary arch.

## 1. INTRODUCTION

Facial esthetics is an inherently subjective discipline. The facial midline and dental midline play important role in facial esthetics, cosmetic dentistry, facial plastic surgery, and anthropologic studies.[1, 2] Thus, dento-facial esthetics helps to produce esthetic beauty and smile facial. The smile esthetics can be grouped into four sections: facial esthetics, gingival esthetics, macroesthetics, and microesthetics.[3] The facial midline forms an important part of esthetic smile design, and it is recommended to flush the dental and facial midline in orthodontic and cosmetic restorative procedures.[4]

Facial and dental midlines greatly vary, and their deviation is seen among the people. Dentists and non-dental personnel can notice the deviations of facial and dental midlines. It is found that the higher the deviations, the more it is easier to detect.[4] Midline deviations more than 2 mm can be detected easily compared to 1-2 mm and ~1 mm. Furthermore, the location of the facial and dental midlines also depends on the clinician.[4] Facial and dental midlines can be located and examined clinically in patients or in 2D photographs.[5, 6] Various reference points on the face can be used to locate the midline and examine the midline deviations.[7] The dental-facial midlines have not been fully investigated and there is no standard classification of midline deviation. Hence, this study aimed to study the face and dental midline and to classify the dental-facial midline using Nepalese subjects.

## 2. MATERIALS AND METHODS

This cross-sectional study was done on 150 Nepalese subjects (55 males and 95 females) from October 2017 till April 2019. Subjects were selected using the following criteria: (1) age of the participants: 18-30 years old, both males and females; (2) not treated with any facial surgery involving the face; and (3) obvious problems that could disfigure of the face. The details of the study subjects [Table 1]. After obtaining the ethical approval from the Institutional Review Committee of the Kathmandu University School of Medical Sciences (IRC-KUSMS) (29/16), the participants recruited in this study and confidentiality of the information was maintained.

**Table 1.** Details of the study subjects in the study.

| Subjects details | Frequency |
| --- | --- |
| **Total subjects** | 150 |
| • **Male** | 80 (53.33%) |
| • **Female** | 70 (46.67%) |
| **Mean age** | 26.5 years |
| **Range** | 18 – 35 years |

All patients read the instructions carefully and they were examined. The facial and dental midlines were located and analyzed using a scale. The facial midline, which divides the face into a right half and left half, was taken as the line passing the glabella, nose tip, the midpoint of the philtrum, and middle of the chin.[8] Maxillary midline is taken the line passing between the upper central incisors and the mandibular midline is taken the line passing between the lower central incisors.[8] All the examination was performed by one examiner. The occurrence with an explanation of the face and dental midline of the patients was as follows (Figure 1 and Figure 2).

**Fig. (1).** The diagrammatic representation of classification of the dental-facial midline. **Class I**. The facial midline is coinciding with both maxillary and mandibular dental midline (I). **Class II**. The facial midline is coinciding with only the maxillary dental midline. The mandibular midline is shifted to the right side (IIa). The mandibular midline is shifted to the left side (IIb). **Class III**. The facial midline is coinciding with only the mandibular dental midline. The maxillary midline is shifted to the right side ((IIIa). The maxillary midline is shifted to the left side (IIIb). **Class IV**. The facial midline is not coinciding with both maxillary and mandibular dental midline. The maxillary and mandibular midline shifted to the right side (IVa). The maxillary and mandibular midline shifted to the left side (IVb). The maxillary midline shifted to the right side and mandibular midline shifted to the left side (IVc). The maxillary midline shifted to the left side and mandibular midline shifted to the right side (IVd). **Class V**. Other than those in the Class I, II, III or IV.

**Fig. (2).** The classification of the dental-facial midline.

Analysis of the data was done using SPSS version 18 for Windows. The comparison of midline was done using Chai-squared Test at a 95% confidence interval.

## 3. RESULTS

Table **1** shows the details of the study subject and the mean age of the subjects was 26.5 years old (range 18-35 years old). It showed only in 16% (24 subjects), the facial midline was coinciding with both the upper and lower midline, as shown in Fig. **1** and Fig. **2**. Females (13, 54.16%) showed greater coincidence of facial and dental midlines than males (11, 45.83%). It showed that dental midline discrepancy was seen more in the maxillary arch compared to the mandibular arch and seen more on the right side. It showed that the majority of the subjects showed a discrepancy of 1 mm, followed by 2 mm, and 3 mm.

Table (**2**) shows the results of the dental-facial midline in the study. It showed that only in 24 (16%) subjects, the facial midline was coinciding with the both the maxillary and the mandibular dental midlines. In 44 (30%) subjects, the facial midline was coinciding with the maxillary dental midline only and in 26 (17%) subjects, the facial midline was coinciding with the mandibular dental midline only. The majority of the upper and lower dental midlines were located on the right side compared to the left side. In 56 (37%) subjects, the facial midline was not coinciding with both upper and lower dental midline. There was no significant difference between tamong the groups in class II-IV (P value <0.0001).

**Table 2.**
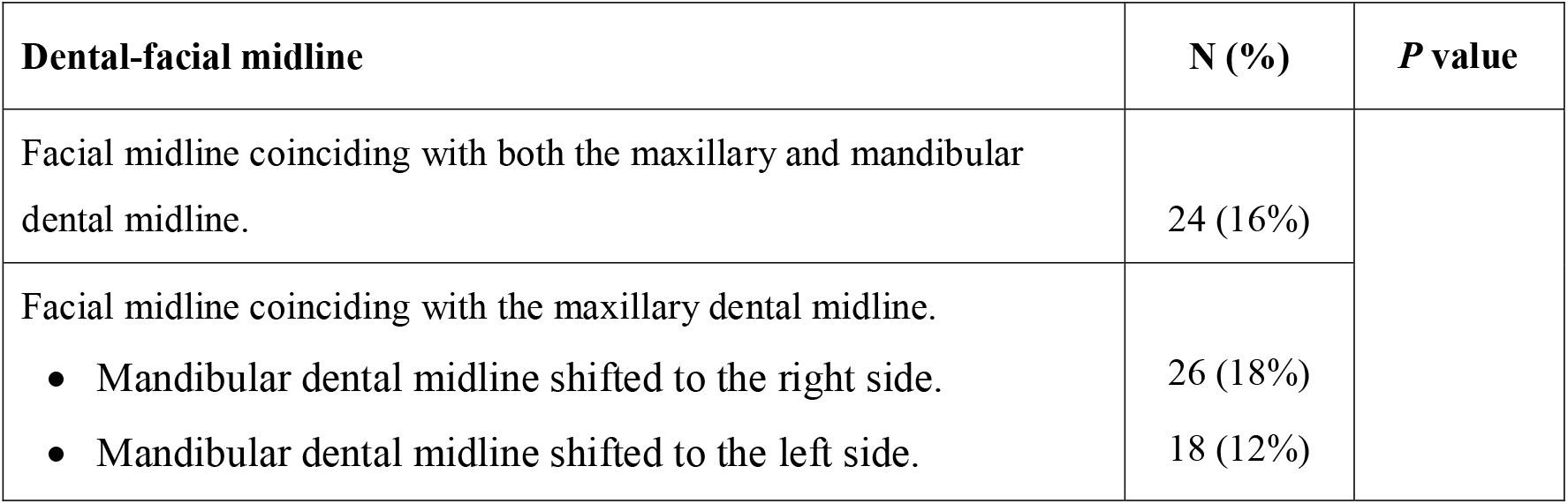
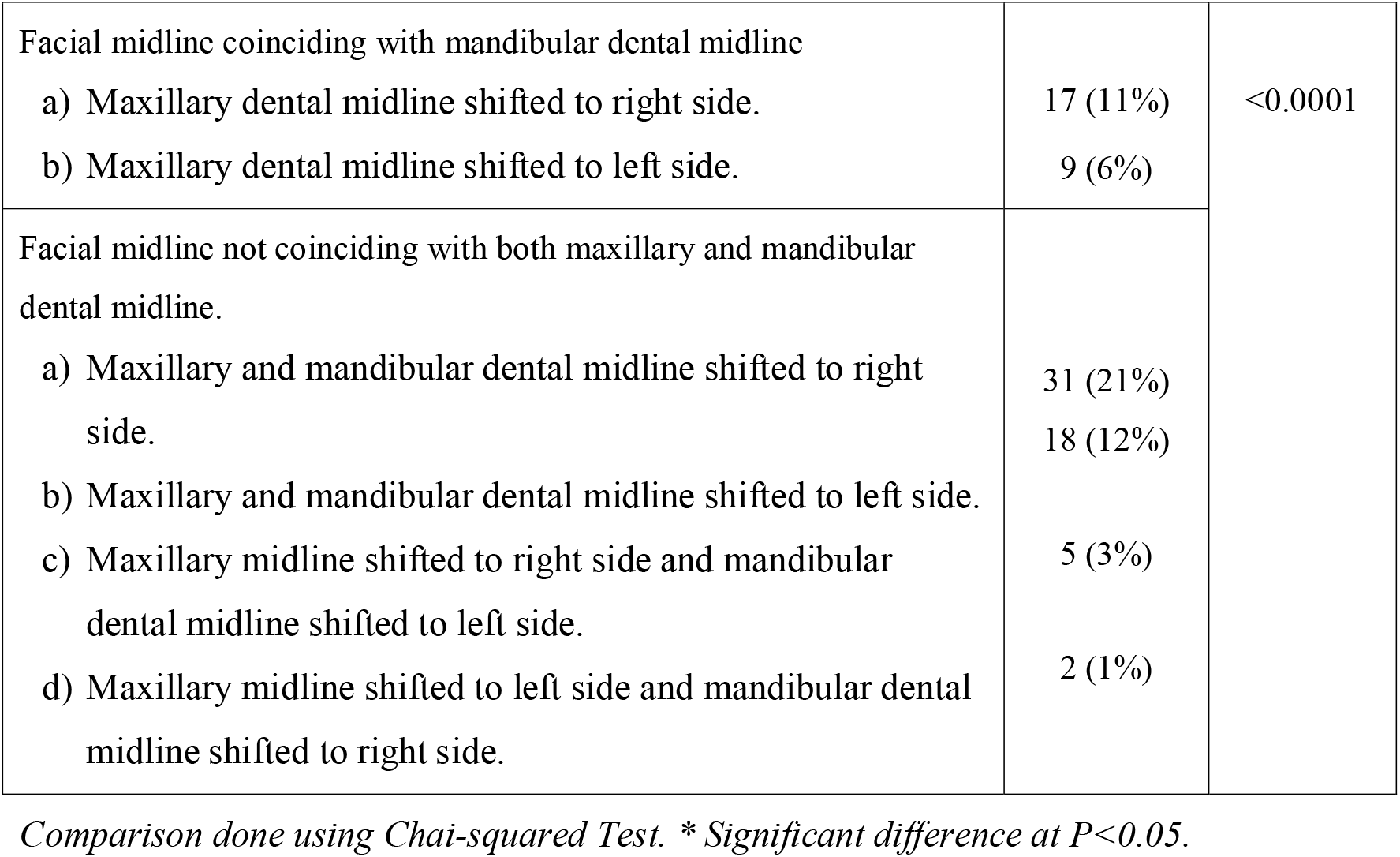
Results of the dental-facial midline in the study.

## 4. DISCUSSION

The facial and dental midlines have an essential role in facial esthetics and facial beauty.[9–11] The dental midline deviation with gender also affects young people’s esthetic perception.[12] The facial anatomic landmarks are important for measuring the facial and dental midlines discrepancy.[13] Besides, 2D photographs can be also used to study the dental midline discrepancy from reference points.[7] Still, the clinical assessment of anatomical landmarks and midline is a practical method to identify the facial and dental midlines and quantify discrepancies. Hence, in this study, we assessed the dental-facial midline clinically.

The results of the midline in our study are similar to the results obtained from the study done by the Mille et al. [5] where they found that the upper and lower midlines were not coinciding in almost 3/4^th^ of the studied subjects. Similarly, Janson et al. [14] mentioned that a small deviation (2 mm) in the dental midline is acceptable by both dentists and orthodontists and patients. But a small axial midline angulation (10°) is already easily noticeable. Furthermore, Khan et al. [15] studied the dento-facial midline in Pakistani subjects and they found that the upper midline was coinciding with lower midline only in 2/3^rd^ of the subjects and facial midline was coinciding with the dental midlines in <50% of the subjects. But they found that dental midline deviations were more on the left side which was contrast to our study. Similarly, a study was done in India on dental-facial midline found that only 20% of the subjects show upper and lower midlines coincidence.[6] Besides, a deviation of 0–1 mm was seen more in girls (55%) than boys (45%), a deviation of 1–2 mm was seen more in boys (54%) than girls (33%), and a deviation of 2–3 mm was seen more in boys (37%) than girls (8%). Similarly, in our study, the coincidence of facial and dental midlines was slightly greater in females than males.

In this study, the participants who participated in this study were subjects from different parts of the country. One limitation of this study is that this study was done on only 150 subjects in Nepal. This study can be done in a larger population and may also be applied in other Asian subjects to find the most common type of dental-facial midline in Asia.

## 5. CONCLUSIONS

The dental-facial midline plays an important parameter for facial and dental esthetics. It was found that most of the subjects show a mild discrepancy of dental-facial midline and the coincidence of the dental and facial midlines is uncommon. Females show a slightly greater coincidence of the facial and dental midline compared to males. The discrepancy of midline was more prevalent in the upper arch and more prevalent on the right side. Mild dental-facial midline discrepancy (1-2 mm) is acceptable and most often may not be perceived.

## Data Availability

The data that support the findings of this study are available from the corresponding author, [D. R.], upon reasonable request.

## ETHICS APPROVAL AND CONSENT TO PARTICIPATE

The study received ethical approval from the Institutional Review Committee of the Kathmandu University School of Medical Sciences (IRC-KUSMS),

## CONSENT FOR PUBLICATION

All participants in the study agreed for the research and study publication.

---

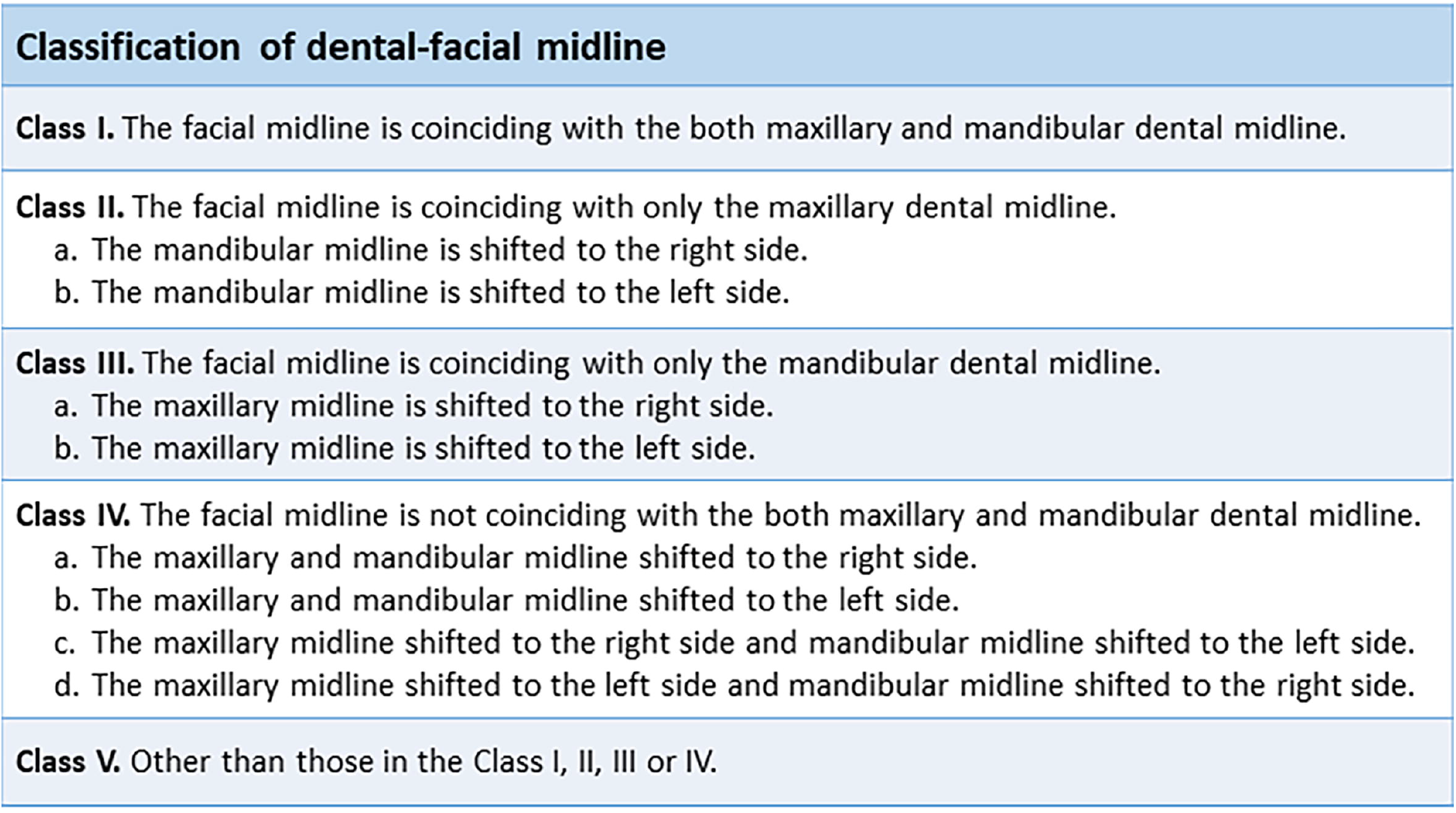

